# Prevalence and source analysis of COVID-19 misinformation of 138 countries

**DOI:** 10.1101/2021.05.08.21256879

**Authors:** Md. Sayeed Al-Zaman

**Affiliations:** Department of Journalism and Media Studies, Jahangirnagar University, Savar, Dhaka, Bangladesh

**Keywords:** COVID-19 misinformation, social media, Facebook, Twitter

## Abstract

This study analyzed 9,657 pieces of misinformation that originated in 138 countries and fact-checked by 94 organizations. Collected from Poynter Institute’s official website and following a quantitative content analysis method along with descriptive statistical analysis, this research produces some novel insights regarding COVID-19 misinformation. The findings show that India (15.94%), the US (9.74%), Brazil (8.57%), and Spain (8.03%) are the four most misinformation-affected countries. Based on the results, it is presumed that the prevalence of COVID-19 misinformation can have a positive association with the COVID-19 situation. Social media (84.94%) produces the highest amount of misinformation, and the internet (90.5%) as a whole is responsible for most of the COVID-19 misinformation. Moreover, Facebook alone produces 66.87% misinformation among all social media platforms. Of all countries, India (18.07%) produced the highest amount of social media misinformation, perhaps thanks to the country’s higher internet penetration rate, increasing social media consumption, and users’ lack of internet literacy. On the other hand, countries like Turkey, the US, Brazil, and the Philippines where either political control over media is intense or political conservatism is apparent, experienced a higher amount of misinformation from mainstream media, political figures, and celebrities. Although the prevalence of misinformation was the highest in March 2020, given the present trends, it may likely to increase slightly in 2021.

## 1. Introduction

This study endeavored to understand the prevalence and source of COVID-19 misinformation around the world. From 2020 until now, a search in scholarly databases (e.g., Scopus and Web of Science) shows that more than 500 research investigated COVID-19 misinformation from different disciplines, such as communication, psychology, politics, and medical sciences (1–9). Apart from disciplinary and thematic analysis, many of such studies investigated the sources of misinformation and misinformation scenarios in different countries as well (4,9–11). However, these studies are limited in one or more of the following ways: (a) they selected either only one or a very few countries for their analysis of misinformation; (b) their sample of misinformation cases were smaller compared to its higher prevalence; (c) insights on misinformation in different countries according to different sources were unavailable; (b) comparative source analysis of misinformation was limited. Therefore, considering these limitations, in the present study, we analyzed 9,657 pieces of misinformation originated from 138 countries and debunked by 94 fact-checking organizations to understand the frequency of misinformation in different countries, sources, and country-wise sources distributions. The results suggest that the prevalence of misinformation does not follow any geographical pattern, rather it might be consistent with the pandemic casualty-led tension and information vacuum. Also, social media poses a big challenge to public health and health communication by producing most of the COVID-19 misinformation. The following discussion is divided into three main sections. In the next section, the details of data collection and analysis have been described.

## 2. Methodology

This exploratory study sought to understand the prevalence and sources of COVID-19 misinformation around the world. Three specific inquiries of this study were:

*RQ1*: Which countries are most affected by COVID-19 misinformation?
*RQ2*: What sources produce most of the COVID-19 misinformation?
*RQ3*: What sources of misinformation are dominant in which countries?

The data for this study were collected from the official website of Poynter Institute for Media Studies (https://www.poynter.org/ifcn-covid-19-misinformation). It is a Florida-based non-profit organization established in 1975, which is actively working on reducing the prevalence of misinformation around the world. The organization has two specialized branches dedicated to fact-checking: PolitiFact and the International Fact-Checking Network (IFCN). From the beginning of the COVID-19 pandemic, Poynter Institute has been combating COVID-19 misinformation and collecting prevalent and popular misinformation cases from different countries, and including them on their website. We collected these data using Web Scraper, an automated scraping extension for web browsers (see more http://webscraper.io). In this automated web scraping, we extracted the claims of misinformation, fact-checkers, sources, dates, countries, types of claims, and explanations of misinformation. We limited our study period from 1 January 2020 to 1 March 2021, although March 2021 only included a single day, i.e., 1 March 2021. During this period, the website included 9,657 COVID-19 misinformation from 94 IFCN-certified fact-checkers. Important to note that the data we collected were publicly available. Such public data are not subject to copyright and can be utilized for research purposes (12). Also, unlike semi-public and private data, our data did not require *informed consent* (13). Moreover, in recent academic scholarship, ethical scraping for research purposes is permissible (14). Previous studies also used similar web scrapers to collect research data (e.g., Hautea, Parks, Takahashi, and Zeng, 2021). Therefore, the data we collected and used in this study were free from ethical obligations.

After collecting the data, at first, we cleaned them by eliminating the unnecessary and fragmented parts. This phase also included language corrections and style corrections. Finally, we chose two variables for our analysis that were relevant to our research questions: country name and misinformation source. The dataset mainly included 9,518 pieces of misinformation from 138 countries, and country names for 139 more misinformation were missing. For RQ1, we calculated the percentage of each country to understand their contributions to the total share of misinformation. We also calculated both aggregate and country-wise monthly misinformation distributions to show the changes in misinformation counts throughout the period. For RQ2, we coded the misinformation sources as follows: social media, mainstream media, popular bodies, other internet sources, and miscellaneous. Previous studies identified online media and mainstream media as the two major media sources of COVID-19 misinformation (4). In this study, however, we divided online media into two types: social media and other internet sources. We further categorized social media and mainstream media according to their media type (e.g., Facebook, Twitter, television, newspaper). In this process, we found that some social media and mainstream media were not specified so that we coded them accordingly. Regarding the category *popular bodies*, we observed that, in many cases, political figures and organizations, and celebrities have been playing roles in producing and circulating misinformation. Therefore, we introduced it as a new category. This category specifically included four main sources: political parties, groups, and figures (e.g., president); non-political organizations (e.g., religious organization); celebrities, influencers, and popular non-political figures (e.g., film actor); relevant specialists and responsible persons (e.g., doctor). We also observed that except for social media, COVID-19 misinformation had a few more internet sources: we categorized them as online portal and blog. The online portals included online news portals and other websites. Finally, we analyzed the countries and their sources of misinformation using cross-tabulation. Consistent with the research questions, instead of inferential statistics, this study adopted descriptive statistical analyses including frequency and percentage analysis and cross-tabulations to analyze and interpret the data. For data preparation and analysis, we used Microsoft Excel 2019 and IBM SPSS Statistics 25. For data visualization, we used Tableau 2020.4, an AI-powered professional data visualization and analysis software.

## 3. Results & Discussion

The present study aimed at answering three questions on COVID-19 misinformation: most affected countries, most popular sources, and popularity of sources according to the countries. A content analysis along with descriptive statistics analyzed 9,657 pieces of misinformation from 138 countries to answer the research inquiries. To the best of our knowledge, this study dealt with the largest amount of COVID-19 misinformation from the highest number of countries and sources to date.

The map shows that a few countries in the world suffered from the higher pervasiveness of misinformation (Figure 1). Most of the Asian and African countries had a lower amount of misinformation, while South Asia, and North and South America show a higher amount of misinformation. India had the highest amount of misinformation during the period (*n* = 1,691; 15.94%), followed by the United States (*n* = 1,032; 9.74%), and Brazil (*n* = 909; 8.57%) (Table 1). The top four countries include a European country as well, i.e., Spain (*n* = 852; 8.03%). These four countries had comparatively higher amounts of COVID-19 misinformation than other countries. For example, Columbia is in the fifth position with 400 (3.77%) misinformation, which is less than half of Spain’s frequency. This result shows that COVID-19 misinformation is not concentrated in some specific geographic areas, rather it is decentralized all over the world. Also, a few countries experience more misinformation than most countries. It seems that misinformation in the top countries is somewhat consistent with the casualties they experienced during the pandemic. That means the prevalence of misinformation may have a positive association with pandemic-led casualties. For example, in both the list of COVID-19 misinformation and COVID-19 casualties, the following ten countries are common among the top fifteen countries: India, the United States (US) Brazil, Spain, France, Turkey, Columbia, Argentina, Italy, and Mexico (16). Since the present paper was unable to establish this correlation, we would like to invite more research to explain this presumption empirically.

**Table 1.** Countries and their counts of COVID-19 misinformation.

| Rank | Country | Frequency | Percentage | Rank | Country | Frequency | Percentage |
| --- | --- | --- | --- | --- | --- | --- | --- |
| 1 | India | 1691 | 15.94% | 71 | Algeria | 10 | 0.09% |
| 2 | United States | 1032 | 9.74% | 72 | Kyrgyzstan | 10 | 0.09% |
| 3 | Brazil | 909 | 8.57% | 73 | Mali | 10 | 0.09% |
| 4 | Spain | 852 | 8.03% | 74 | Singapore | 10 | 0.09% |
| 5 | Colombia | 400 | 3.77% | 75 | Burundi | 9 | 0.08% |
| 6 | France | 356 | 3.36% | 76 | Madagascar | 9 | 0.08% |
| 7 | Philippines | 339 | 3.20% | 77 | Saudi Arabia | 9 | 0.08% |
| 8 | Ukraine | 300 | 2.83% | 78 | Syria | 9 | 0.08% |
| 9 | Turkey | 288 | 2.72% | 79 | Chile | 8 | 0.08% |
| 10 | Mexico | 250 | 2.36% | 80 | Ethiopia | 8 | 0.08% |
| 11 | Argentina | 235 | 2.22% | 81 | Ivory Coast | 8 | 0.08% |
| 12 | Italy | 231 | 2.18% | 82 | Sweden | 8 | 0.08% |
| 13 | Taiwan | 223 | 2.10% | 83 | Belarus | 7 | 0.07% |
| 14 | Georgia | 212 | 2.00% | 84 | Cameroon | 7 | 0.07% |
| 15 | North Macedonia | 167 | 1.57% | 85 | Europe | 7 | 0.07% |
| 16 | Missing | 139 | 1.31% | 86 | Iraq | 7 | 0.07% |
| 17 | Australia | 131 | 1.24% | 87 | Senegal | 7 | 0.07% |
| 18 | Germany | 129 | 1.22% | 88 | Austria | 5 | 0.05% |
| 19 | Sri Lanka | 119 | 1.12% | 89 | Cuba | 5 | 0.05% |
| 20 | Middle East | 116 | 1.09% | 90 | United Arab Emirates | 5 | 0.05% |
| 21 | Indonesia | 115 | 1.08% | 91 | Lebanon | 4 | 0.04% |
| 22 | Kenya | 115 | 1.08% | 92 | South Sudan | 4 | 0.04% |
| 23 | Nigeria | 106 | 1.00% | 93 | Southern Africa | 4 | 0.04% |
| 24 | North Africa | 101 | 0.95% | 94 | Bulgaria | 3 | 0.03% |
| 25 | Ecuador | 95 | 0.90% | 95 | El Salvador | 3 | 0.03% |
| 26 | Canada | 92 | 0.87% | 96 | Gabon | 3 | 0.03% |
| 27 | Portugal | 89 | 0.84% | 97 | Libya | 3 | 0.03% |
| 28 | Poland | 84 | 0.79% | 98 | Montenegro | 3 | 0.03% |
| 29 | Ireland | 83 | 0.78% | 99 | Romania | 3 | 0.03% |
| 30 | Croatia | 78 | 0.74% | 100 | Uruguay | 3 | 0.03% |
| 31 | Bolivia | 71 | 0.67% | 101 | Zimbabwe | 3 | 0.03% |
| 32 | United Kingdom | 70 | 0.66% | 102 | Afghanistan | 2 | 0.02% |
| 33 | Myanmar | 65 | 0.61% | 103 | Asia | 2 | 0.02% |
| 34 | Greece | 64 | 0.60% | 104 | Azerbaijan | 2 | 0.02% |
| 35 | Venezuela | 64 | 0.60% | 105 | Czech Republic | 2 | 0.02% |
| 36 | Japan | 58 | 0.55% | 106 | Finland | 2 | 0.02% |
| 37 | Lithuania | 56 | 0.53% | 107 | Guinea | 2 | 0.02% |
| 38 | Ghana | 45 | 0.42% | 108 | Iceland | 2 | 0.02% |
| 39 | Russia | 43 | 0.41% | 109 | Kuwait | 2 | 0.02% |
| 40 | South Africa | 43 | 0.41% | 110 | North America | 2 | 0.02% |
| 41 | Egypt | 42 | 0.40% | 111 | Norway | 2 | 0.02% |
| 42 | Thailand | 40 | 0.38% | 112 | South America | 2 | 0.02% |
| 43 | Costa Rica | 39 | 0.37% | 113 | Switzerland | 2 | 0.02% |
| 44 | Bosnia and<br>Herzegovina | 37 | 0.35% | 114 | Yemen | 2 | 0.02% |
| 45 | Belgium | 35 | 0.33% | 115 | Albania | 1 | 0.01% |
| 46 | Hong Kong | 35 | 0.33% | 116 | Bahrain | 1 | 0.01% |
| 47 | Serbia | 35 | 0.33% | 117 | Burkina Faso | 1 | 0.01% |
| 48 | China | 33 | 0.31% | 118 | Central America | 1 | 0.01% |
| 49 | Kazakhstan | 31 | 0.29% | 119 | Dominican<br>Republic | 1 | 0.01% |
| 50 | Peru | 31 | 0.29% | 120 | Fiji | 1 | 0.01% |
| 51 | Netherlands | 27 | 0.25% | 121 | Gambia | 1 | 0.01% |
| 52 | South Korea | 25 | 0.24% | 122 | Honduras | 1 | 0.01% |
| 53 | Latvia | 23 | 0.22% | 123 | Hungary | 1 | 0.01% |
| 54 | Pakistan | 22 | 0.21% | 124 | Kosovo | 1 | 0.01% |
| 55 | Paraguay | 21 | 0.20% | 125 | Malawi | 1 | 0.01% |
| 56 | Tunisia | 21 | 0.20% | 126 | Moldova | 1 | 0.01% |
| 57 | Western Sahara | 19 | 0.18% | 127 | North Korea | 1 | 0.01% |
| 58 | Jordan | 17 | 0.16% | 128 | Oman | 1 | 0.01% |
| 59 | Uganda | 17 | 0.16% | 129 | Papua-New-Guinea | 1 | 0.01% |
| 60 | Denmark | 16 | 0.15% | 130 | Qatar | 1 | 0.01% |
| 61 | East Africa | 16 | 0.15% | 131 | Rwanda | 1 | 0.01% |
| 62 | New Zealand | 16 | 0.15% | 132 | Slovakia | 1 | 0.01% |
| 63 | Tanzania | 16 | 0.15% | 133 | Sudan | 1 | 0.01% |
| 64 | Africa | 15 | 0.14% | 134 | Tajikistan | 1 | 0.01% |
| 65 | Israel | 15 | 0.14% | 135 | Timor Leste | 1 | 0.01% |
| 66 | Guatemala | 14 | 0.13% | 136 | Tuvalu | 1 | 0.01% |
| 67 | Malaysia | 13 | 0.12% | 137 | Uzbekistan | 1 | 0.01% |
| 68 | Congo | 11 | 0.10% | 138 | Vanuatu | 1 | 0.01% |
| 69 | Iran | 11 | 0.10% | 139 | Vietnam | 1 | 0.01% |
| 70 | Morocco | 11 | 0.10% |  |  |  |  |

**Figure 1.**
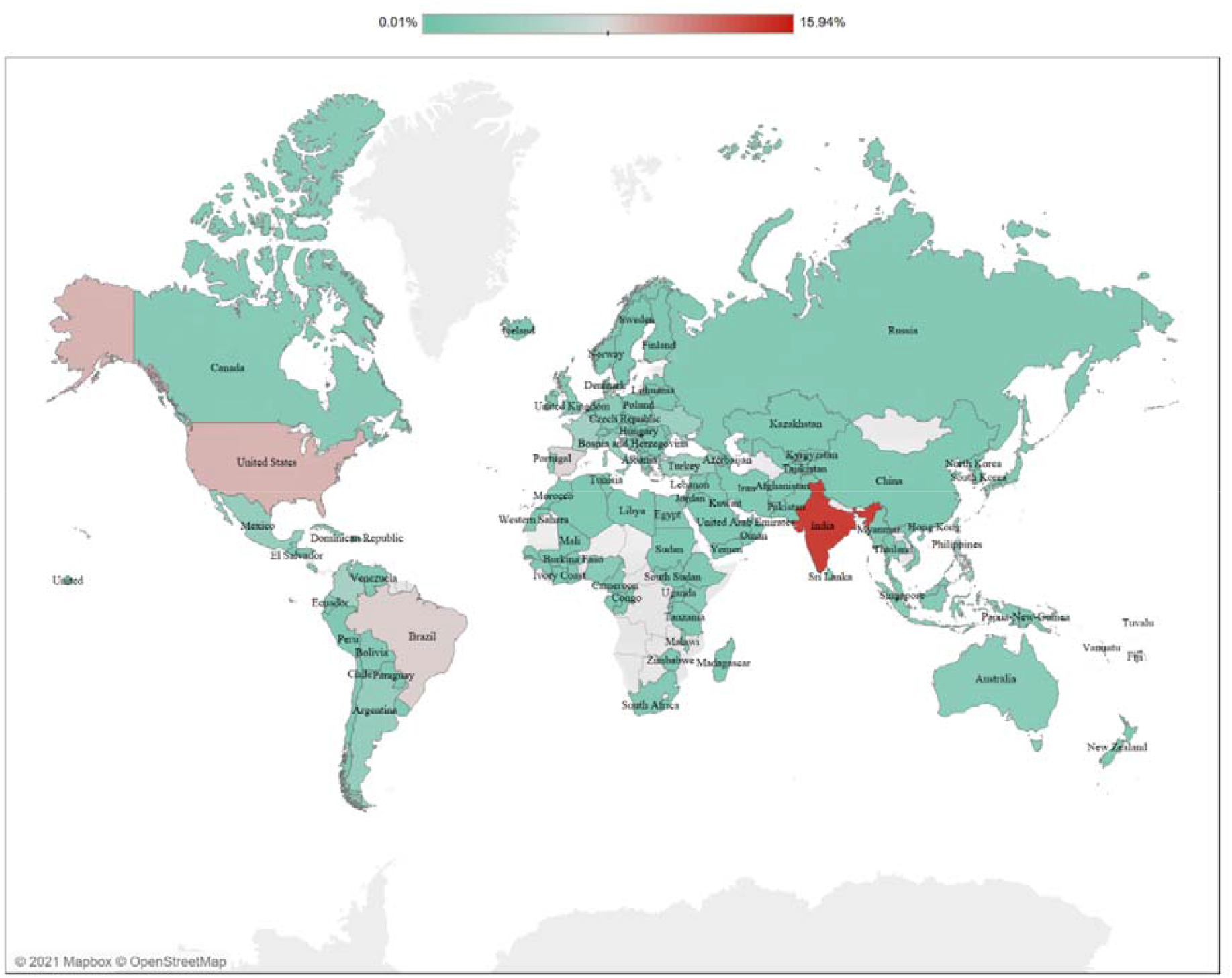
Percentage map of the COVID-19 misinformation.

The aggregate amount of misinformation is showing a gradual decline after May 2020 with some infrequent and small surges (Figure 2). Misinformation reached its peak in March 2020 (*n* = 9,256) that started dropping in the subsequent months: 8,416, 4,772, and 2,320 in April, May, and June, respectively. The number reached 752 in February 2021. The trend also shows a downward tendency. However, a forecast shows that the number may likely to increase and remain to 955 pieces of misinformation on average in March-September 2021. Most of the countries including India, the US, Brazil, and Spain experienced a surge in misinformation towards March and July 2020 (Table 2). However, a few countries, such as Georgia, experienced a surge from September to December 2020. Both the aggregated and country-wise misinformation count hint that the prevalence of misinformation could be consistent with the number of casualties. Put another way, misinformation surged before or amid the infection, and death rates surged. For example, from March 2020, India, the US, Brazil, and Spain were experiencing a gradual rise in COVID-19 cases along with COVID-19 misinformation (16). On the other hand, before September, Georgia had almost no or limited COVID-19 cases along with COVID-19 misinformation (17). This result is consistent with the hypothesis of Difonzo and Bordia (2006) who claimed that misinformation has a positive correlation with tension, damages, and information scarcity. However, as stated earlier, since this study could not perform a correlation coefficient analysis between the amount of misinformation and the number of COVID-19 casualties, the hypothesis would remain as subject to further analysis. Previous studies did not analyze the relationship either. A few studies only investigated users’ engagement with COVID-19 contents. For instance, a study analyzed Twitter conversations on COVID-19 from January to March 2020 and observed the gradual increase in misinformation with time (7). Another study explored that misinformation also increased with time like general conversations and reached the peak in March 2020 (10). Users’ engagement with COVID-19 content may increase or decrease based on a few factors, such as negative news on COVID-19, misinformation, and vaccine news (19).

**Table 2.**
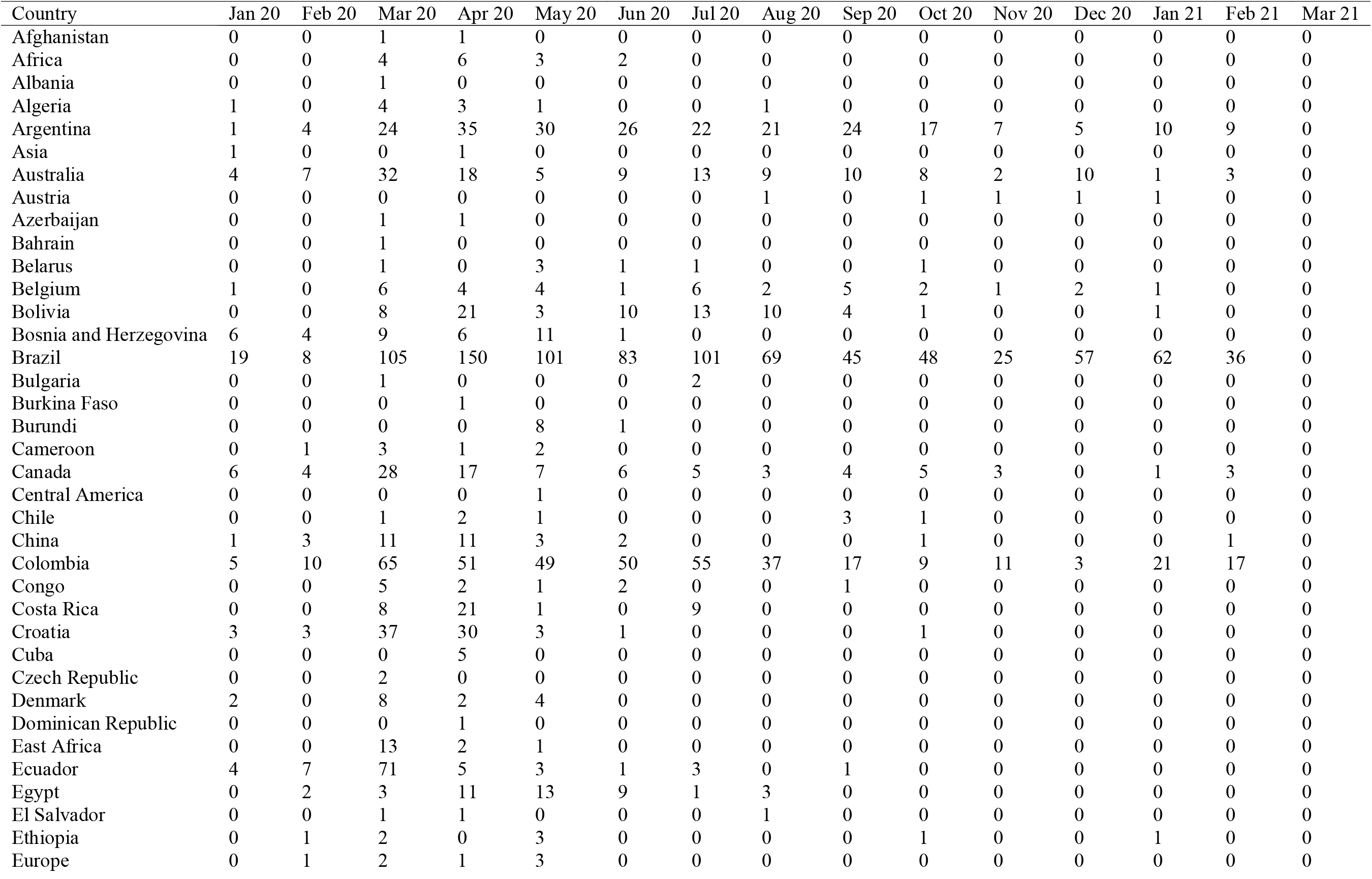
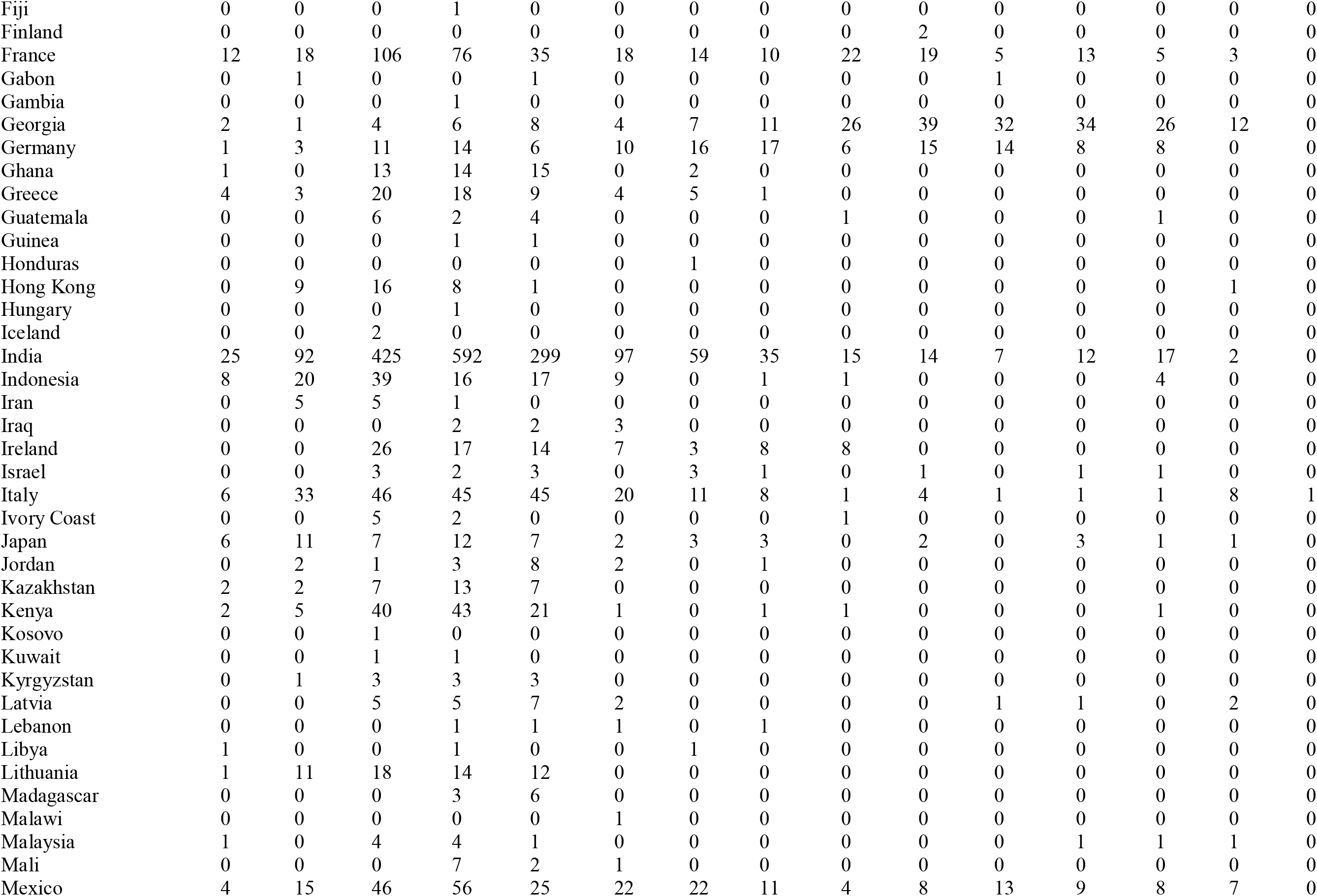
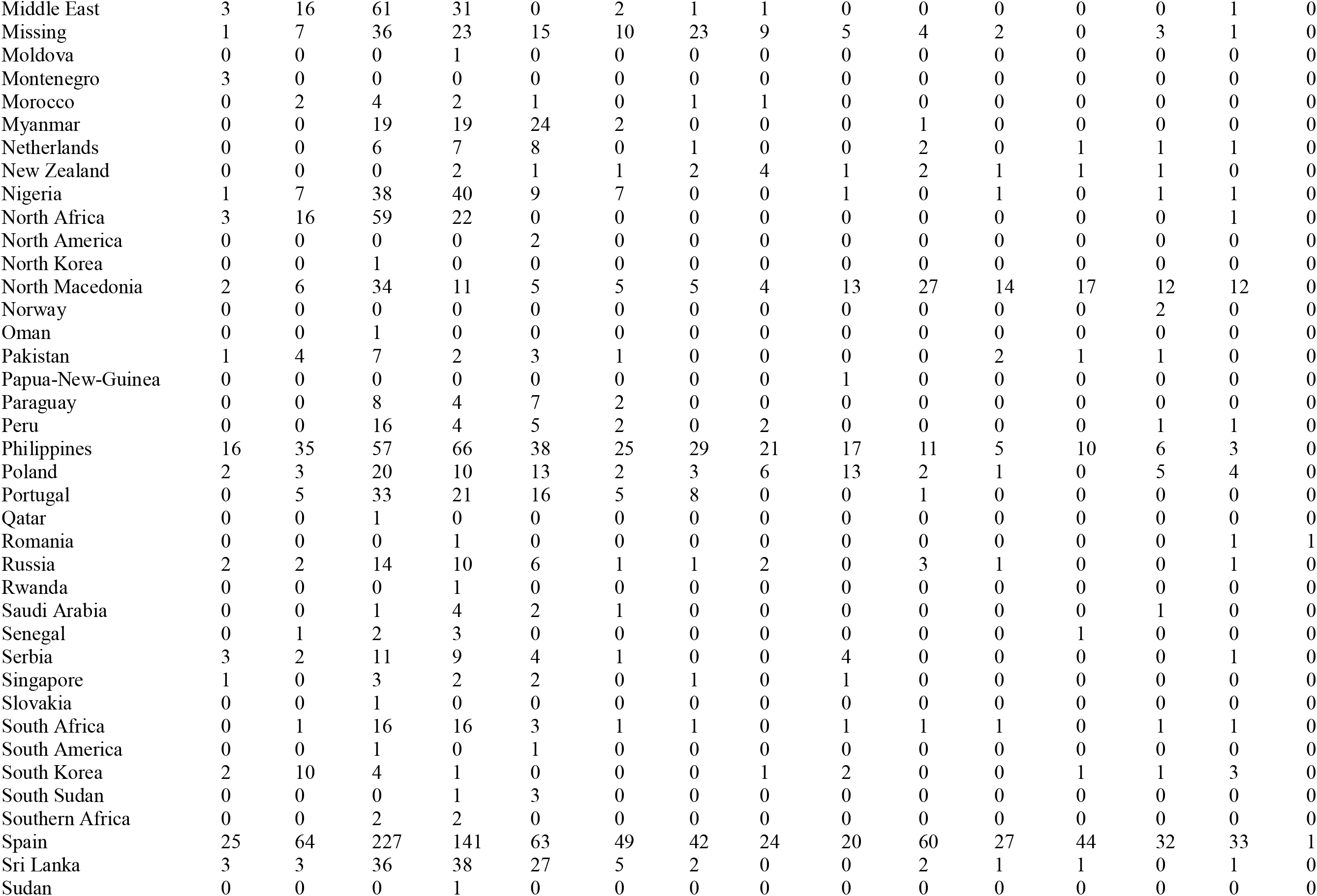
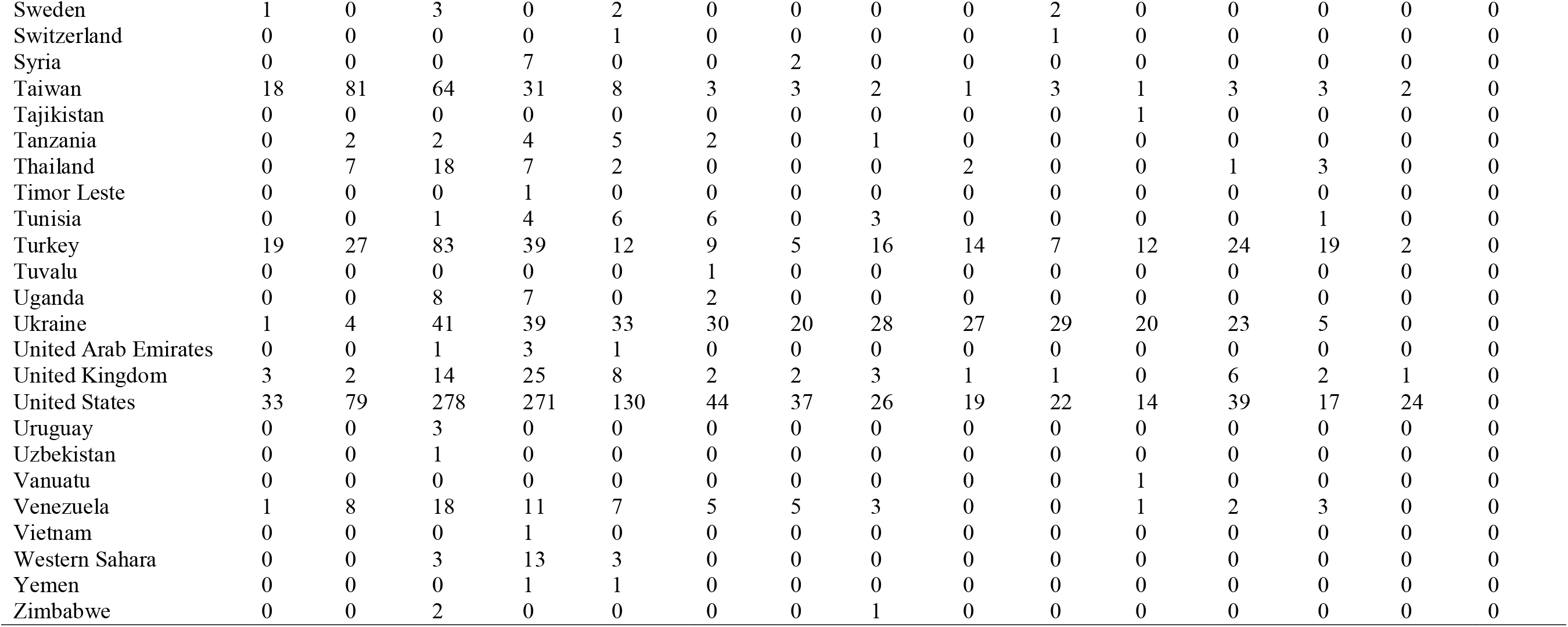
Monthly frequency distributions of misinformation according to the countries.

**Table 3.**
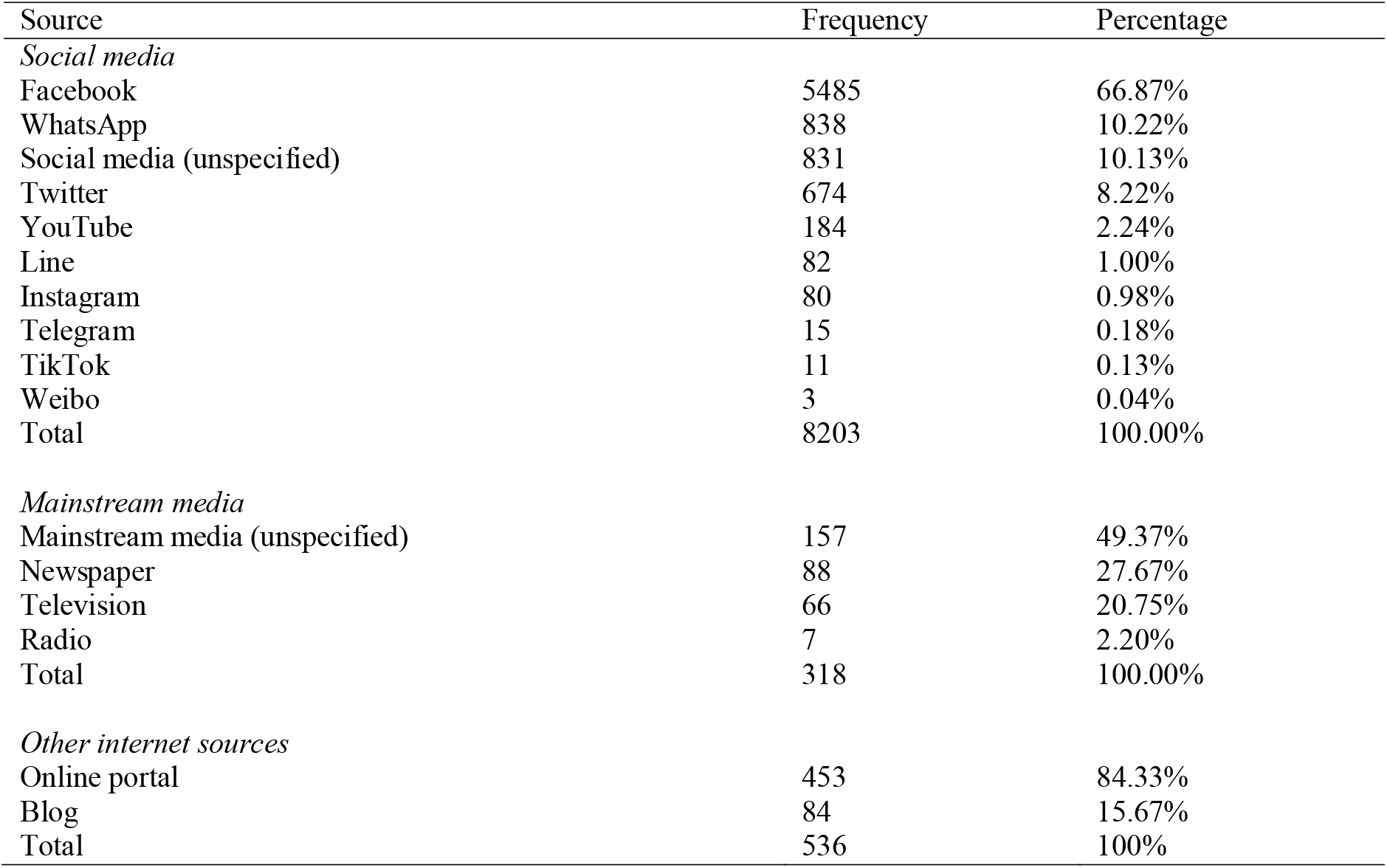
Various social media, mainstream media, and internet sources of misinformation.

| Source | Frequency | Percentage |
| --- | --- | --- |
| *Social media* |  |  |
| Facebook | 5485 | 66.87% |
| WhatsApp | 838 | 10.22% |
| Social media (unspecified) | 831 | 10.13% |
| Twitter | 674 | 8.22% |
| YouTube | 184 | 2.24% |
| Line | 82 | 1.00% |
| Instagram | 80 | 0.98% |
| Telegram | 15 | 0.18% |
| TikTok | 11 | 0.13% |
| Weibo | 3 | 0.04% |
| Total | 8203 | 100.00% |
| *Mainstream media* |  |  |
| Mainstream media (unspecified) | 157 | 49.37% |
| Newspaper | 88 | 27.67% |
| Television | 66 | 20.75% |
| Radio | 7 | 2.20% |
| Total | 318 | 100.00% |
| *Other internet sources* |  |  |
| Online portal | 453 | 84.33% |
| Blog | 84 | 15.67% |
| Total | 536 | 100% |

**Table 4.** Sources of misinformation according to the countries.

| Country | Source of misinformation (%) |  |  |  |  |
| --- | --- | --- | --- | --- | --- |
|  | Social media | Mainstream media | Popular bodies | Other internet sources | Misc. |
| Afghanistan | 0.02% | 0.00% | 0.00% | 0.00% | 0.00% |
| Africa | 0.17% | 0.00% | 0.00% | 0.00% | 0.00% |
| Albania | 0.01% | 0.00% | 0.00% | 0.00% | 0.00% |
| Algeria | 0.09% | 0.00% | 0.00% | 0.16% | 0.47% |
| Argentina | 2.30% | 2.24% | 2.56% | 0.81% | 1.89% |
| Asia | 0.01% | 0.00% | 0.23% | 0.00% | 0.00% |
| Australia | 1.42% | 0.00% | 0.23% | 0.32% | 0.00% |
| Austria | 0.04% | 0.00% | 0.00% | 0.16% | 0.00% |
| Azerbaijan | 0.01% | 0.00% | 0.00% | 0.16% | 0.00% |
| Bahrain | 0.01% | 0.00% | 0.00% | 0.00% | 0.00% |
| Belarus | 0.07% | 0.00% | 0.00% | 0.16% | 0.00% |
| Belgium | 0.31% | 0.28% | 0.47% | 0.49% | 0.47% |
| Bolivia | 0.47% | 0.56% | 1.63% | 0.16% | 8.96% |
| Bosnia and Herzegovina | 0.09% | 3.08% | 0.23% | 2.59% | 0.47% |
| Brazil | 9.17% | 1.40% | 9.77% | 5.83% | 0.94% |
| Bulgaria | 0.01% | 0.28% | 0.23% | 0.00% | 0.00% |
| Burkina Faso | 0.01% | 0.00% | 0.00% | 0.00% | 0.00% |
| Burundi | 0.10% | 0.00% | 0.00% | 0.00% | 0.00% |
| Cameroon | 0.07% | 0.00% | 0.00% | 0.16% | 0.00% |
| Canada | 0.79% | 0.84% | 1.40% | 0.97% | 2.83% |
| Central America | 0.01% | 0.00% | 0.00% | 0.00% | 0.00% |
| Chile | 0.09% | 0.00% | 0.00% | 0.00% | 0.00% |
| China | 0.36% | 0.00% | 0.00% | 0.16% | 0.00% |
| Colombia | 3.83% | 2.80% | 3.02% | 4.85% | 1.42% |
| Congo | 0.12% | 0.00% | 0.00% | 0.00% | 0.00% |
| Costa Rica | 0.42% | 0.00% | 0.23% | 0.00% | 0.00% |
| Croatia | 0.55% | 4.76% | 0.00% | 1.78% | 0.47% |
| Cuba | 0.03% | 0.28% | 0.00% | 0.16% | 0.00% |
| Czech Republic | 0.02% | 0.00% | 0.00% | 0.00% | 0.00% |
| Denmark | 0.16% | 0.00% | 0.00% | 0.16% | 0.47% |
| Dominican Republic | 0.01% | 0.00% | 0.00% | 0.00% | 0.00% |
| East Africa | 0.13% | 0.84% | 0.23% | 0.00% | 0.00% |
| Ecuador | 1.05% | 0.00% | 0.00% | 0.16% | 0.00% |
| Egypt | 0.47% | 0.00% | 0.00% | 0.00% | 0.00% |
| El Salvador | 0.01% | 0.00% | 0.23% | 0.00% | 0.47% |
| Ethiopia | 0.07% | 0.00% | 0.00% | 0.32% | 0.00% |
| Europe | 0.06% | 0.28% | 0.00% | 0.16% | 0.00% |
| Fiji | 0.01% | 0.00% | 0.00% | 0.00% | 0.00% |
| Finland | 0.02% | 0.00% | 0.00% | 0.00% | 0.00% |
| France | 3.18% | 1.96% | 6.28% | 4.69% | 3.30% |
| Gabon | 0.02% | 0.00% | 0.00% | 0.16% | 0.00% |
| Gambia | 0.01% | 0.00% | 0.00% | 0.00% | 0.00% |
| Georgia | 1.87% | 8.40% | 0.47% | 1.94% | 0.00% |
| Germany | 1.16% | 0.28% | 0.47% | 3.56% | 0.00% |
| Ghana | 0.39% | 0.00% | 0.00% | 1.62% | 0.00% |
| Greece | 0.17% | 7.84% | 0.23% | 2.91% | 0.94% |
| Guatemala | 0.12% | 0.00% | 0.47% | 0.16% | 0.00% |
| Guinea | 0.02% | 0.00% | 0.00% | 0.00% | 0.00% |
| Honduras | 0.01% | 0.00% | 0.00% | 0.00% | 0.00% |
| Hong Kong | 0.37% | 0.00% | 0.23% | 0.00% | 0.47% |
| Hungary | 0.01% | 0.00% | 0.00% | 0.00% | 0.00% |
| Iceland | 0.02% | 0.00% | 0.00% | 0.00% | 0.00% |
| India | 18.07% | 8.12% | 3.95% | 2.75% | 1.89% |
| Indonesia | 1.07% | 0.28% | 1.40% | 1.94% | 0.00% |
| Iran | 0.08% | 0.28% | 0.47% | 0.00% | 0.47% |
| Iraq | 0.08% | 0.00% | 0.00% | 0.00% | 0.00% |
| Ireland | 0.90% | 0.28% | 0.00% | 0.16% | 0.00% |
| Israel | 0.13% | 0.84% | 0.00% | 0.00% | 0.00% |
| Italy | 2.11% | 3.36% | 1.40% | 3.72% | 0.00% |
| Ivory Coast | 0.09% | 0.00% | 0.00% | 0.00% | 0.00% |
| Japan | 0.50% | 0.28% | 0.47% | 1.13% | 1.42% |
| Jordan | 0.19% | 0.00% | 0.00% | 0.00% | 0.00% |
| Kazakhstan | 0.17% | 0.28% | 0.47% | 1.94% | 0.47% |
| Kenya | 0.98% | 2.24% | 0.47% | 2.59% | 0.47% |
| Kosovo | 0.01% | 0.00% | 0.00% | 0.00% | 0.00% |
| Kuwait | 0.01% | 0.00% | 0.00% | 0.16% | 0.00% |
| Kyrgyzstan | 0.04% | 0.28% | 0.23% | 0.65% | 0.00% |
| Latvia | 0.18% | 0.00% | 1.40% | 0.16% | 0.00% |
| Lebanon | 0.04% | 0.00% | 0.00% | 0.00% | 0.00% |
| Libya | 0.03% | 0.00% | 0.00% | 0.00% | 0.00% |
| Lithuania | 0.50% | 0.56% | 0.70% | 0.16% | 2.36% |
| Madagascar | 0.09% | 0.00% | 0.23% | 0.00% | 0.00% |
| Malawi | 0.01% | 0.00% | 0.00% | 0.00% | 0.00% |
| Malaysia | 0.14% | 0.00% | 0.00% | 0.00% | 0.00% |
| Mali | 0.11% | 0.00% | 0.00% | 0.00% | 0.00% |
| Mexico | 2.66% | 1.12% | 0.23% | 0.65% | 0.94% |
| Middle East | 1.29% | 0.00% | 0.00% | 0.00% | 0.00% |
| Missing | 0.77% | 1.12% | 1.16% | 0.00% | 28.77% |
| Moldova | 0.01% | 0.00% | 0.00% | 0.00% | 0.00% |
| Montenegro | 0.01% | 0.56% | 0.00% | 0.00% | 0.00% |
| Morocco | 0.10% | 0.00% | 0.00% | 0.32% | 0.00% |
| Myanmar | 0.71% | 0.00% | 0.00% | 0.16% | 0.00% |
| Netherlands | 0.18% | 0.56% | 0.70% | 0.81% | 0.47% |
| New Zealand | 0.18% | 0.00% | 0.00% | 0.00% | 0.00% |
| Nigeria | 1.00% | 0.56% | 0.23% | 1.78% | 0.94% |
| North Africa | 1.12% | 0.00% | 0.00% | 0.00% | 0.00% |
| North America | 0.02% | 0.00% | 0.00% | 0.00% | 0.00% |
| North Korea | 0.00% | 0.00% | 0.00% | 0.16% | 0.00% |
| North Macedonia | 1.20% | 2.52% | 0.70% | 7.12% | 1.42% |
| Norway | 0.02% | 0.00% | 0.00% | 0.00% | 0.00% |
| Oman | 0.01% | 0.00% | 0.00% | 0.00% | 0.00% |
| Pakistan | 0.22% | 0.00% | 0.00% | 0.00% | 0.94% |
| Papua-New-Guinea | 0.01% | 0.00% | 0.00% | 0.00% | 0.00% |
| Paraguay | 0.17% | 0.56% | 0.93% | 0.00% | 0.00% |
| Peru | 0.29% | 0.00% | 1.16% | 0.00% | 0.00% |
| Philippines | 2.98% | 0.56% | 8.60% | 4.69% | 1.42% |
| Poland | 0.69% | 0.56% | 0.23% | 3.07% | 0.00% |
| Portugal | 0.87% | 0.00% | 0.23% | 0.97% | 1.89% |
| Qatar | 0.01% | 0.00% | 0.00% | 0.00% | 0.00% |
| Romania | 0.02% | 0.00% | 0.23% | 0.00% | 0.00% |
| Russia | 0.31% | 0.28% | 0.47% | 1.94% | 0.00% |
| Rwanda | 0.01% | 0.00% | 0.00% | 0.00% | 0.00% |
| Saudi Arabia | 0.10% | 0.00% | 0.00% | 0.00% | 0.00% |
| Senegal | 0.08% | 0.00% | 0.00% | 0.00% | 0.00% |
| Serbia | 0.11% | 2.24% | 1.63% | 1.62% | 0.00% |
| Singapore | 0.11% | 0.00% | 0.00% | 0.00% | 0.00% |
| Slovakia | 0.01% | 0.00% | 0.00% | 0.00% | 0.00% |
| South Africa | 0.46% | 0.00% | 0.23% | 0.16% | 0.00% |
| South America | 0.02% | 0.00% | 0.00% | 0.00% | 0.00% |
| South Korea | 0.24% | 0.56% | 0.23% | 0.00% | 0.00% |
| South Sudan | 0.03% | 0.00% | 0.00% | 0.00% | 0.47% |
| Southern Africa | 0.04% | 0.00% | 0.00% | 0.00% | 0.00% |
| Spain | 7.83% | 6.72% | 6.98% | 6.63% | 25.00% |
| Sri Lanka | 1.29% | 0.00% | 0.00% | 0.49% | 0.00% |
| Sudan | 0.01% | 0.00% | 0.00% | 0.00% | 0.00% |
| Sweden | 0.06% | 0.28% | 0.23% | 0.16% | 0.00% |
| Switzerland | 0.02% | 0.00% | 0.00% | 0.00% | 0.00% |
| Syria | 0.10% | 0.00% | 0.00% | 0.00% | 0.00% |
| Taiwan | 2.38% | 0.56% | 0.00% | 0.49% | 1.89% |
| Tajikistan | 0.01% | 0.00% | 0.00% | 0.00% | 0.00% |
| Tanzania | 0.11% | 0.00% | 0.00% | 0.97% | 0.00% |
| Thailand | 0.42% | 0.00% | 0.00% | 0.00% | 0.94% |
| Timor Leste | 0.01% | 0.00% | 0.00% | 0.00% | 0.00% |
| Tunisia | 0.22% | 0.00% | 0.00% | 0.16% | 0.00% |
| Turkey | 2.48% | 12.32% | 0.70% | 2.91% | 0.00% |
| Tuvalu | 0.01% | 0.00% | 0.00% | 0.00% | 0.00% |
| Uganda | 0.14% | 0.00% | 0.23% | 0.49% | 0.00% |
| Ukraine | 3.15% | 2.80% | 0.47% | 0.65% | 0.47% |
| United Arab Emirates | 0.06% | 0.00% | 0.00% | 0.00% | 0.00% |
| United Kingdom | 0.69% | 1.12% | 0.23% | 0.49% | 0.00% |
| United States | 8.61% | 11.20% | 31.16% | 12.94% | 2.36% |
| Uruguay | 0.03% | 0.00% | 0.00% | 0.00% | 0.00% |
| Uzbekistan | 0.01% | 0.00% | 0.00% | 0.00% | 0.00% |
| Vanuatu | 0.01% | 0.00% | 0.00% | 0.00% | 0.00% |
| Venezuela | 0.50% | 0.56% | 3.26% | 0.00% | 1.42% |
| Vietnam | 0.00% | 0.28% | 0.00% | 0.00% | 0.00% |
| Western Sahara | 0.21% | 0.00% | 0.00% | 0.00% | 0.00% |
| Yemen | 0.02% | 0.00% | 0.00% | 0.00% | 0.00% |
| Zimbabwe | 0.03% | 0.00% | 0.00% | 0.00% | 0.00% |

**Figure 2.**
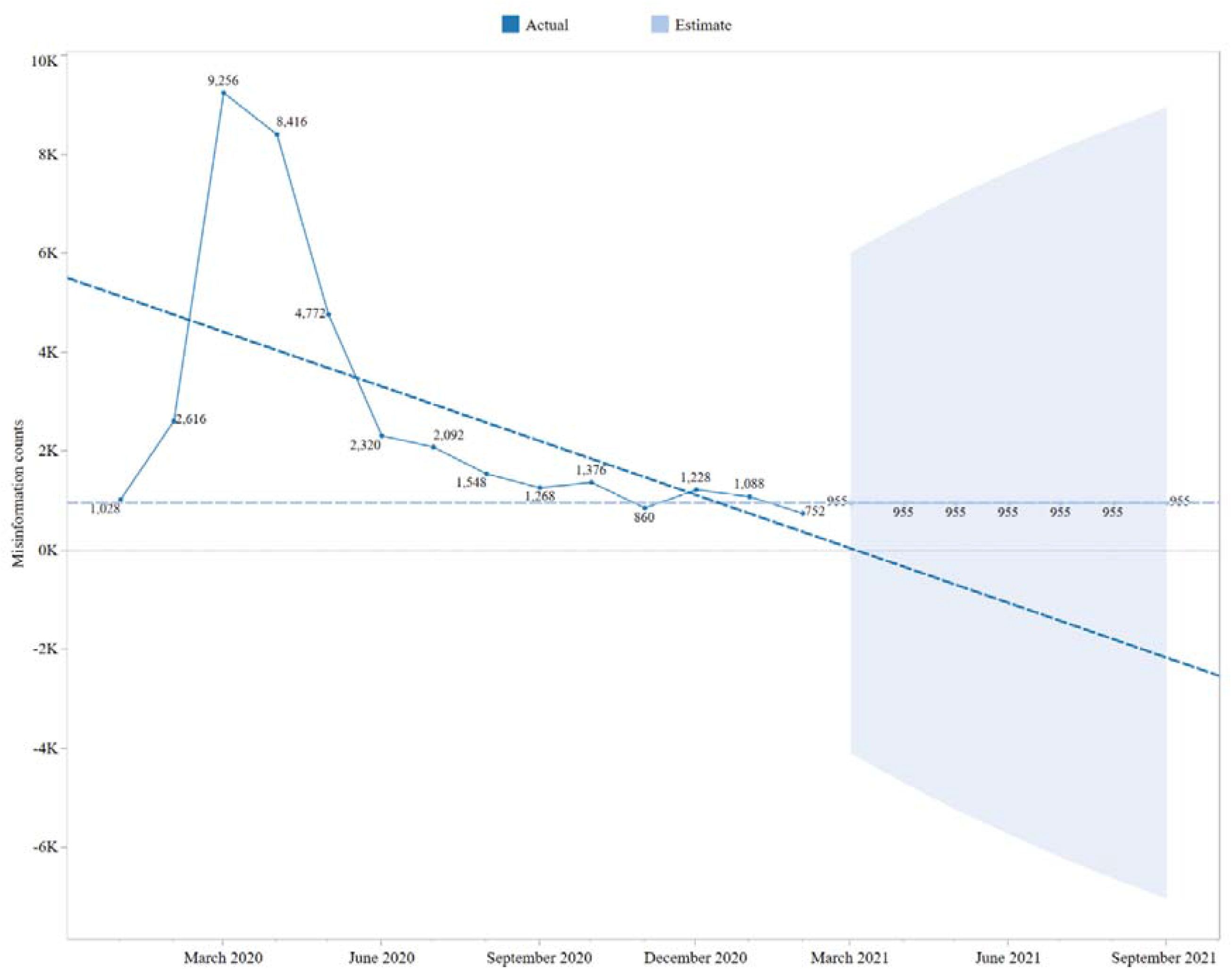
Misinformation trends during the period and a trend-based forecast.

Of the four main sources, social media produced the highest amount of misinformation (84.94%), followed by other internet sources (5.56%) (Figure 3). Interestingly, the internet-based sources alone produced 90.5% of all misinformation. It suggests that the internet is the ultimate producer of COVID-19 misinformation, which requires further attention from both scholars and policymakers of the respected countries where internet-based misinformation is more prevalent. On the contrary, the share of mainstream media in misinformation production was relatively lower (3.29%) than the other sources. Misinformation from all sources was consistent from January to December 2020 with a few smaller surges. For example, social media starting from 82.61% in January reached 88.93% in December experiencing at least four minor decreases in February, May, July, and November. All sources except other internet sources reached their highest in March 2020: social media reached 23.82%, mainstream media to 28.62%, and popular bodies to 21.16%. Only other internet sources reached the peak (22.16%) in April. A survey shows that 63.3% of respondents encounter most of the COVID-19 misinformation in social media than other sources (11). Previous studies focusing on different regions also revealed that despite benefiting the public by providing useful information, social media is producing profuse COVID-19 misinformation (1,8,11,20,21). For that reason, social media is addressed as a “double-edged sword” and social media misinformation is addressed as “an [misinformation] epidemic within the COVID-19 pandemic” (8,21).

**Figure 3.**
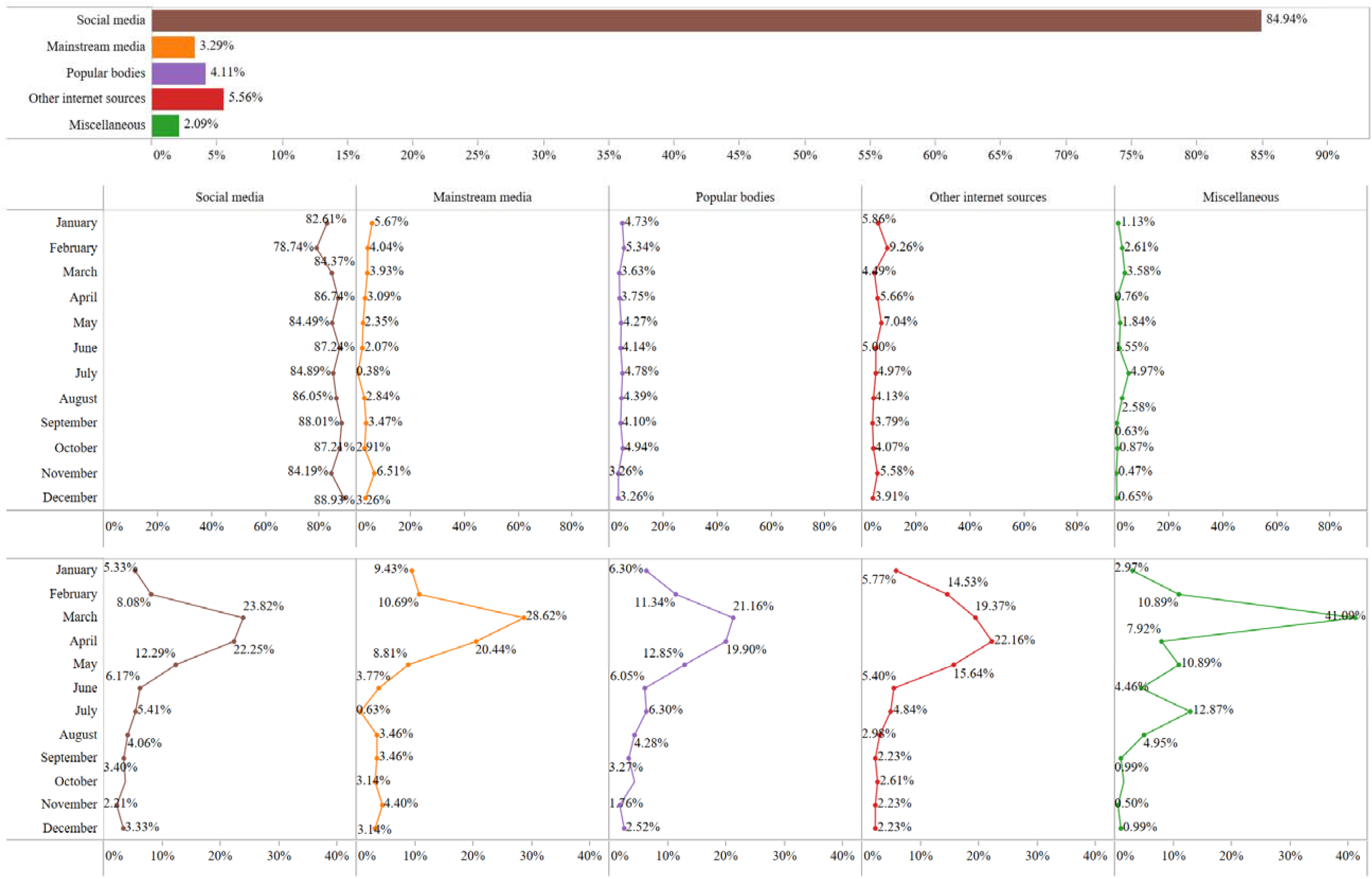
Details of the COVID-19 misinformation sources.

In social media, Facebook was the most prominent source of misinformation that alone produced 66.87% (*n* = 5,485) of the total social media misinformation. WhatsApp, a popular instant messaging application, is in the second position with only 10.22% (*n* = 838) misinformation. Twitter is in the third position on the list with 8.22% (*n* = 674) misinformation. Previous studies also explored that these three social media platforms are more responsible for COVID-19 misinformation propagation (1,4,10,22). Approximately 831 (10.13%) pieces of social media misinformation and 157 (49.37%) pieces of mainstream media misinformation had no specific platform mentioned. In mainstream media, newspapers (*n* = 88; 27.67%) produced a higher amount of misinformation than television channels (*n* = 66; 20.75%), meaning print media produces more misinformation than broadcast media. The online portal is also an important source of misinformation (*n* = 453) that produced a higher amount of misinformation than mainstream media (*n* = 318) and a few popular social media platforms like YouTube (*n* = 184), Line (*n* = 82), and Instagram (*n* = 80). Of online portals, most of them were news portals, containing unreliable information regarding COVID-19. For example, Asembi News, one of Ghana’s biggest news websites, published the following false claim: “Family of three died within days of each other before testing positive for coronavirus.”

Of all countries, India produced the highest amount of social media misinformation (18.07%), followed by Brazil (9.17%) and the US (8.61%). The reason for India’s social media misinformation epidemic could be: (a) the higher social media penetration rates from the last few years, which may increase further in the next few years (23–25); (b) the increased consumption of social media contents during the pandemic (26); (c) social media users’ lack of digital literacy that makes them the victims of misinformation (27). On the other hand, Turkey produced the highest amount of mainstream media misinformation (12.32%), followed by the US (11.20%) and Georgia (8.40%). Lack of press freedom, authoritarian control over mainstream media, and the government-endorsed disinformation campaign using the media might be responsible for Turkey’s higher mainstream media misinformation (28,29). Interestingly, the US alone produced 31.16% misinformation from popular bodies, mostly from the political figures, groups, and celebrities, which is unprecedented. Brazil (9.77%) and the Philippines (8.60%) followed the US in this respect. Some political and sociocultural factors might be responsible for such results. For example, conservative politicians and the political environment is found conducive for COVID-19 misinformation in the US and the public trust on politicians’ approach to tackling COVID-19 was much higher in the country (9,30). Also, in the contemporary popular culture of the US, celebrities have significant influence over social and political events and audiences, which could be another reason for this result. In Brazil, on the other hand, President Bolsonaro himself is a champion of COVID-19 denial, and the government itself produces COVID-19 disinformation (31). Studies also found that political conservatism like these countries is associated with higher susceptibility to misinformation (9). Like the previous category, i.e., popular bodies, the US produced the highest amount of misinformation from different internet sources as well (12.94%), followed by North Macedonia (7.12%) and Spain (6.63%). For India, the highest misinformation-producing country, the amount of misinformation from popular bodies (3.95%) and various internet sources (2.75) was moderate.

The higher prevalence of misinformation would complicate public health responses and health communication in many countries. Meanwhile, COVID-19 misinformation had claimed many lives around the world (1,32). In countries like India and Bangladesh, religious and political COVID-19 misinformation is propelling interreligious discontents and encouraging superstitions and unscientific health practices (4,33–35). Therefore, proper measures should be sanctioned to control the prevalence of misinformation to reduce health hazards.

## 4. Conclusion

To conclude, this study produced a few novel insights regarding COVID-19 misinformation, which would help to better understand the COVID-19 misinformation climate around the world. Also, because this study utilized the largest COVID-19 misinformation data, the results can be more generalizable. Lastly, the scholars may find the results and methodological aspects useful for their future studies. Beyond the contributions and usefulness, however, this study is limited in a few ways. It relied on the data collected by independent fact-checkers, who often have limited resources to collect, research, and debunk all available claims (10). Also, the data included misinformation from only 94 IFCN-approved fact-checkers around the world, which seem insufficient. As a result, many countries were not on the list and popular misinformation from these countries was not included. For example, BD Fact Check, a Bangladeshi fact-checking organization, debunked 300 pieces of misinformation in March-December 2020, and approximately 150 of them were related to the pandemic (36,37), which is missing in the current dataset. It reflects that the amount of COVID-19 misinformation is much higher than that was included in our dataset. Another limitation of the study is that it used only descriptive statistics to observe the variables’ frequencies, percentages, and cross-tabulations, but the inclusion of the results from inferential statistics (e.g., association analysis between the variables) would better explain their relationships. Although it was not a requirement for the present study, future studies may consider it and find it useful.

## Data Availability

N/A

